# Immunogenicity and safety of inactivated whole virion Coronavirus vaccine with CpG (VLA2001) in healthy adults aged 18 to 55: a randomised phase 1 /2 clinical trial

**DOI:** 10.1101/2021.08.13.21262021

**Authors:** Rajeka Lazarus, Christian Taucher, Claire Brown, Irena Corbic, Leon Danon, Katrin Dubischar, Christopher J.A. Duncan, Susanne Eder-Lingelbach, Saul N Faust, Christopher Green, Karishma Gokani, Romana Hochreiter, Johanna Kellett Wright, Dowan Kwon, Alexander Middleditch, Alasdair P.S. Munro, Kush Naker, Florentina Penciu, David Price, Benedicte Querton, Tawassal Riaz, Amy Ross-Russell, Amada Sanchez-Gonzalez, Hayley Wardle, Sarah Warren, Adam Finn, the Valneva Phase 1 Trial Group

## Abstract

**Background:** We assessed the safety, tolerability and immunogenicity of VLA2001 is a whole-virion inactivated SARS-CoV-2 vaccine adsorbed to alum with a toll-like receptor 9 agonist adjuvant in healthy volunteers aged 18-55.

**Methods:** The first 15 participants were enrolled, in groups of 5, to receive two doses, separated by 21 days, of one of three dose concentrations, administered intramuscularly. 138 further participants were randomised 1:1:1 to receive the same 3 dose concentrations, in a double blinded manner. Primary outcomes were solicited adverse reactions 7 days after each vaccination and neutralising antibody geometric mean titres (GMT) against SARS-CoV-2, 2 weeks after the second vaccination (day 36), measured by live microneutralisation assay against wild-type virus (MNA50). Secondary outcomes included unsolicited adverse events, and humoral and cellular responses at day 36, measured by IgG ELISA against Spike protein and interferon-γ secreting T-cells by ELISpot stimulated with multiple SARS-CoV-2 antigens. (ClinicalTrials.gov NCT04671017, ISRCTN 82411169)

**Findings:** Between December 16, 2020 and January 21, 2021, 153 participants were enrolled and randomised evenly between the dose groups. The rates of solicited reactions were similar after the first and second doses and between the three dose groups. The most frequent local reactions were tenderness (58·2%) and pain (41·8%) and systemic reactions were headache (46%) and fatigue (39·2%).

In the high dose group, two weeks following the second dose, the geometric mean titres were 530.4 (95% CI: 421·49, 667·52) for neutralizing antibodies and 2147·9 (95% CI: 1705·98, 2704·22) for S-binding antibodies. There was a dose dependent response with 90·0% (95% CI:78·0%.,97·0%) seroconversion (4-fold rise) at day 36 in the high dose group, which was significantly higher than rates in both the medium (73.5%; 95% CI: 59%,85%), CIs) and low dose (51%; 95%CI: 37%,65%) rate, CIs) groups (both p < 0.001). Antigen-specific interferon-γ T-cells reactive against the S, M and N proteins were observed in 76, 36 and 49% of high dose recipients, respectively.

**Interpretation:** VLA2001-201 was well tolerated and produced both humoral and cellular immune responses, with a clear dose-response effect.

**Funding:** This study was funded by the Department of Health and Social Care, UK

The funder had no role in the study design, implementation or analysis.

## Introduction

The COVID-19 pandemic, caused by the severe acute respiratory syndrome coronavirus 2 (SARS-CoV-2) continues to cause considerable mortality and morbidity worldwide.^1^ Several vaccines against SARS-CoV-2 have been given emergency licenses around the world and millions of individuals have been vaccinated.^2^ Most of these licensed vaccines induce immunity to the SARS-CoV-2 Spike protein which is the viral protein primarily involved in host receptor recognition, attachment and entry into cells.^2^

Inactivated vaccines have been used extensively over past decades to provide protection against a number of pathogens. There are 16 inactivated COVID-19 candidate vaccines in clinical evaluation with some already deployed in many countries and administered to millions of people.^3^ Several studies demonstrate that inactivated vaccines containing aluminium adjuvant induce neutralising antibody responses and have good safety profiles.^4-7^. A desirable characteristic for any COVID-19 vaccine candidate is the ability to induce T-cell responses. Most whole-virion inactivated vaccines are adjuvanted with alum alone, and induce only limited cell-mediated responses.

VLA2001 is a highly purified, inactivated, whole virion, SARS-CoV-2 vaccine, formulated with aluminium hydroxide combined with cytosine phospho-guanine (CpG 1018; Dynavax Technologies Corporation). The immunogenicity of VLA2001 has been demonstrated in murine studies (unpublished data) and was enhanced by the addition of CpG 1018, a toll-like receptor 9 (TLR-9) agonist, stimulating production of innate pro-inflammatory cytokines and a pronounced Th1 immune response. This adjuvant has an established safety profile and is contained in a licensed hepatitis B vaccine.^8^

The aim of the study was to assess safety, reactogenicity and immunogenicity of this novel coronavirus vaccine and to establish the optimal dose to take forward into further clinical development.

## Methods

### Study design and participants

This report describes results from the first 36 days of follow up, including two weeks after a second dose of vaccine, in an ongoing safety and dose-finding immunogenicity trial. Following an initial open-label dose escalation phase, the second phase of the study had a randomised, double-blind parallel group design with a 1:1:1 allocation to three dose levels. Healthy adult volunteers aged 18-55 years were recruited either through local advertisement or a national vaccine volunteer registry. All participants were screened with full medical history and physical examination and blood and urine samples were sent to assess HIV, hepatitis B and C, SARS-CoV2 antibodies, full blood count, clotting function, inflammatory markers, liver and renal function and urinary abnormalities (including blood, protein and glucose). All women of childbearing potential had a serum HCG test performed at screening and urine pregnancy tests at all subsequent visits. Subjects with acute febrile or chronic disease, laboratory-confirmed SARS-CoV2 infection or who participated in other SARS-CoV-2 vaccine clinical studies were excluded; for a full list of exclusion criteria refer to supplementary material.

Written informed consent was obtained from all participants, and the trial was conducted in accordance with the principles of the Declaration of Helsinki and Good Clinical Practice. The study was approved in the UK by the Medicines and Healthcare products Regulatory Agency (43185/0002/001-0001) and the London, Brent ethics committee ref 20/HRA/5205.

### Randomisation and Blinding

The phase 2 study participants were randomly assigned (1:1:1) to receive either the 3AU (low), 7AU (medium) or 35AU (high dose) of VLA2001. The randomisation code was generated by the study statistician and allocation was performed within a secure web platform. The investigational medicinal product (IMP) was provided to study sites in packaging that was identical for all strengths of the vaccine. IMP allocation was based on an identifier linked to the randomisation, therefore as there was no visual difference between the doses, study staff and participants remained blinded to dose allocation.

### Procedures

VLA2001 was produced using a highly purified, whole virus, SARS-CoV-2 strain derived from a Chinese tourist from Hubei, diagnosed with COVID-19 in a hospital in Rome. The virus was cultured in Vero cells and inactivated with β-propiolactone. VLA2001 is adjuvanted with cytosine phospho-guanine (CpG) 1018, a licensed adjuvant, in combination with aluminium hydroxide. The vaccine was manufactured at Valneva, Scotland, according to current Good Manufacturing Practice as described in the Investigational Medicinal Product Dossier and approved by the regulatory agency in the UK. The antigen contents of the vaccines were evaluated using Spike protein ELISA and has been confirmed in the final product to be 3AU, 7AU and 35AU for the low, medium and high doses respectively. The vaccine was given as a 2-dose schedule separated by 21 days. The active substance was combined with CpG 1018 1mg/dose to reach final concentrations in 0.5ml immediately before administration. Vaccines were administered by intramuscular injection into the deltoid.

The initial participants were recruited into an open label, dose escalation study, in December 2020, to receive either the low (n=5), medium (n=5) or high dose (n=5) vaccine. The first participant in each dose group was immunised and then observed for 3 hours and interviewed after 24 hours by telephone, to assess for adverse reactions, before the remaining participants in each dose group were vaccinated sequentially, with each one observed for one hour before the next was vaccinated. These participants were then revaccinated 3 weeks later following the same procedures.

A double-blinded randomised stage was initiated after the safety data from the initial stage of the study, up to 3 days after the last high dose vaccination, had been reviewed by an independent data safety monitoring committee to ensure none of the pre-specified stopping rules had been met.

Participants received a vaccine on Day 0 and Day 21 and were asked to record solicited and unsolicited adverse events using electronic diaries during the 7 days after vaccination. They were provided with a digital thermometer to measure oral temperature every evening from the day of vaccination for 7 days. Solicited local events at the site of vaccination were pain, tenderness, erythema and induration/swelling while systemic events were nausea/vomiting, headache, fatigue and myalgia. Unsolicited events, adverse events of special interest (AESI) and serious adverse events (SAE) were collected up to day 36. AESI were defined as COVID-19 manifestations and complications associated with COVID-19, due to the theoretical risk for disease enhancement, as well as immunemediated disorders due to the addition of the CpG 1018 adjuvant. Changes in laboratory parameters were assessed. All adverse events and abnormal laboratory parameters were graded in accordance with the FDA’s toxicity scale for healthy adult and adolescent volunteers enrolled in preventive vaccine clinical trials.^9^ T-cell responses were classified as reactive if 6 or more spot forming units per 2 × 10^5^ peripheral blood mononuclear cells (PBMC) were present once counts in control cells had been subtracted.

Blood samples were taken on days 0, 8, 21 and 36. Antibody responses at these time points were measured using an immunoassay for IgG to full length S-protein (Nexelis, Canada) and a live microneutralisation assay MNA_50_ against the Victoria strain performed by Public Health England (Porton Down, UK).^10^ Cellular immunity against S-protein, Nucleocapsid protein and Membrane protein was assessed at Oxford Immunotec using T-Spot Discovery SARS-CoV-2 (Oxford, UK).^11^

### Outcomes

The primary outcome was the frequency and severity of solicited adverse events (AE) within 7 days of first or second vaccination. Secondary outcomes in this report were the number and percentage of participants with unsolicited AE related AE, AESI and SAE by severity up to day 36.

The primary immunogenicity outcome was the geometric mean titres (GMT) for SARS-CoV-2 neutralising antibodies at day 36. Secondary immunogenicity outcomes included GMT of Spike protein binding and neutralising antibodies at all time points. T-cell responses (spot forming units per 2 × 10^5^ PBMC) were included as exploratory outcomes. The full list of outcomes is available in the protocol (supplementary material).

### Statistical analysis

This is a descriptive study and formal power calculations were not performed. A total of 150 participants was agreed with regulators to be sufficient for initial safety data. The safety analysis included all participants who received at least a single dose of vaccine, the full analysis set. The immunogenicity analysis was performed on a per-protocol analysis set (PPAS), which excluded participants who received less than two vaccinations, received the wrong study medication or who fulfilled the exclusion criteria. The results for all participants were combined for analysis and reporting including the participants who were enrolled in an open label, non-randomised manner

Differences between treatment groups relating to AEs were assessed for significance using Fisher’s exact test. The number and percentage of subjects with solicited injection site and systemic AEs within 7 days after vaccination along with the exact 95% Clopper-Pearson confidence interval (CI) for all AE rates were presented for each dose group and overall. Differences between the dose groups were assessed for significance using Fisher-Freeman-Halton exact test and p-values were presented for this test. GMTs (CI) were be calculated by taking the antilogarithm of the mean (CI) of the log10 transformed titre. Also, p-values from Kruskal Wallis test were calculated to check if the results were significantly different among dose groups at 5% level of significance. If the test suggested significance, then a pairwise group comparison was perfomed using Dwass, Steel, Critchlow-Fligner (DSCF) multiple comparisons post-hoc procedure to determine which pair of dose groups differed significantly. Secondary immunogenicity analyses include comparison of the SCRs on days 21 and 36 using Fisher-Freeman-Halton exact test. If overall group difference were statistically significant, i.e. p-value for Fisher-Freeman-Halton exact test was ≤ 0.05, then multiplicity adjusted p-values (using Hochberg method) for pairwise group difference were calculated with Fisher’s exact test. Seroconversion was defined as at least a four-fold increase in GMT from baseline. Statistical analysis using SAS^®^ version 9.4 was performed by Valneva and were independently verified by LD using R version 4.0.2.

An independent data and safety monitoring board provided safety oversight. This trial is registered with ClinicalTrials.gov, NCT04671017 and with ISRCTN, 82411169

## Results

Between 16^th^ December 2020 and 21^st^ January 2021 285 individuals aged between 18 and 55 were screened and 153 were enrolled in the study, with 51 participants in each dose group. All randomised participants received the first dose of vaccine but 3 participants did not receive a second dose due to pregnancy, diagnosis of COVID-19 and investigator decision not to administer the second dose following an adverse event (haematuria) considered to be treatment-unrelated, figure 1. The mean age of participants was 33·5 years. 46% were female and 93·5% were white, table 1.

**Figure 1.**
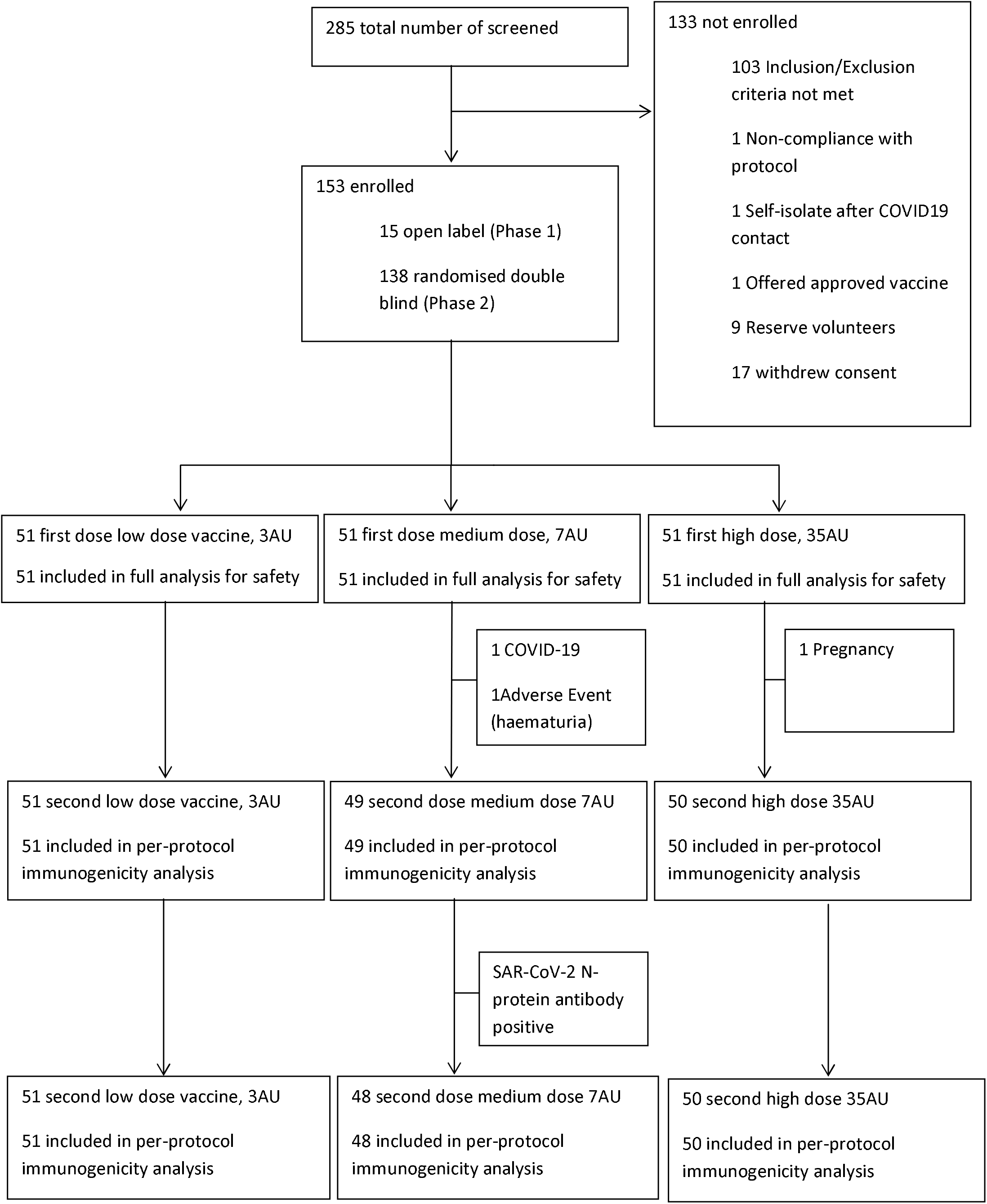
Study Consort Diagram.

**Table 1.**
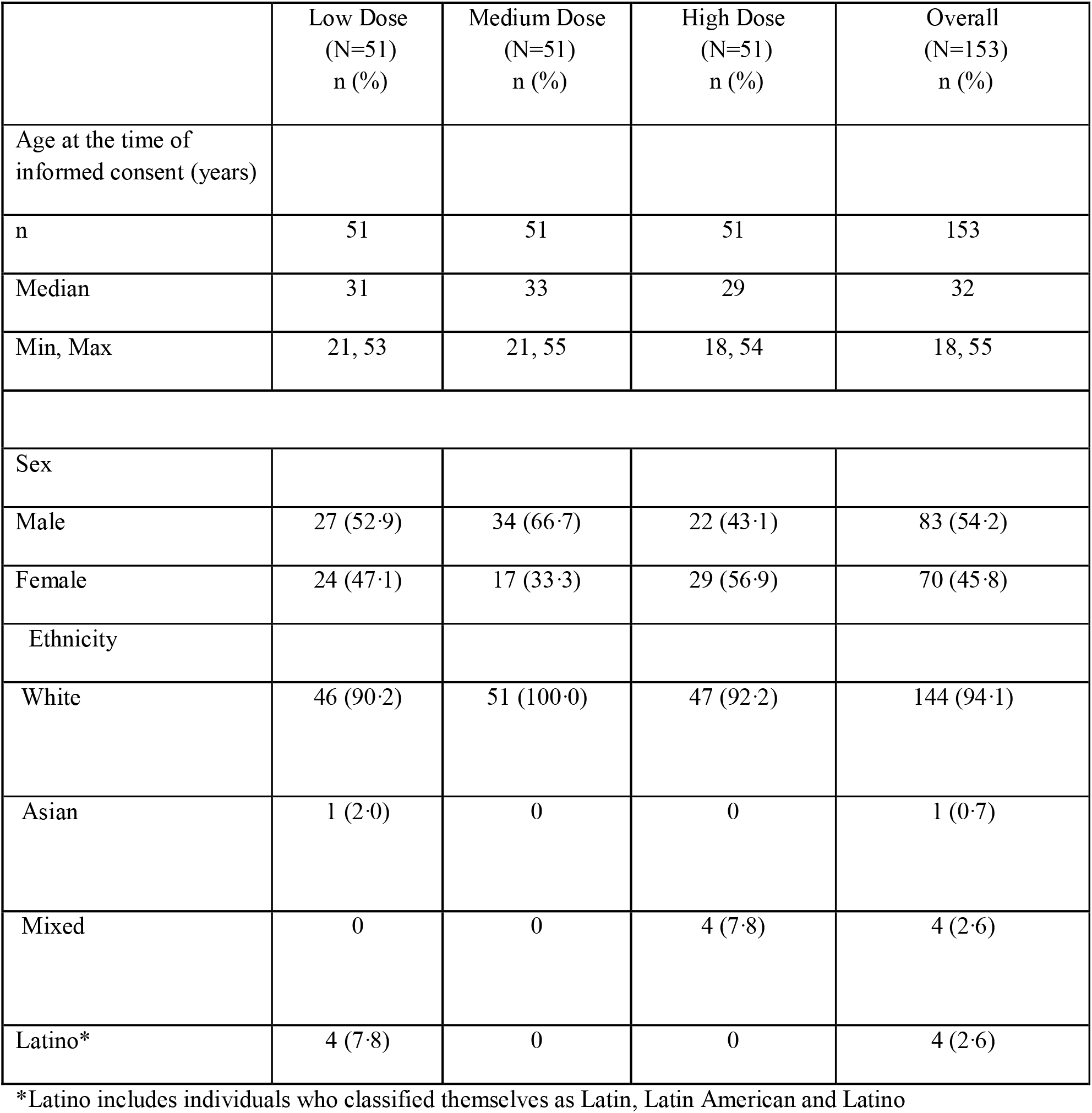
Baseline demographics and characteristics.

|  | Low Dose<br>(N=51)<br>n (%) | Medium Dose<br>(N=51)<br>n (%) | High Dose<br>(N=51)<br>n (%) | Overall<br>(N=153)<br>n (%) |
| --- | --- | --- | --- | --- |
| Age at the time of<br>informed consent (years) |  |  |  |  |
| n | 51 | 51 | 51 | 153 |
| Median | 31 | 33 | 29 | 32 |
| Min, Max | 21, 53 | 21, 55 | 18, 54 | 18, 55 |
| Sex |  |  |  |  |
| Male | 27 (52.9) | 34 (66.7) | 22 (43.1) | 83 (54.2) |
| Female | 24 (47.1) | 17 (33.3) | 29 (56.9) | 70 (45.8) |
| Ethnicity |  |  |  |  |
| White | 46 (90.2) | 51 (100.0) | 47 (92.2) | 144 (94.1) |
| Asian | 1 (2.0) | 0 | 0 | 1 (0.7) |
| Mixed | 0 | 0 | 4 (7.8) | 4 (2.6) |
| Latino* | 4 (7.8) | 0 | 0 | 4 (2.6) |

\*Latino includes individuals who classified themselves as Latin, Latin American and Latino

There were no significant differences in reported reactions between the first and second vaccine doses or the different dose strengths. Overall, 66·7% of vaccinees reported at least one solicited injection site reaction (68·6% low, 60·8% medium and 70·6% high dose groups) with tenderness and injection site pain most commonly reported, table 2a. Overall, 69·3% of vaccinees reported at least one solicited systemic reaction (72·5% low, 62·7% medium and 72·5% high dose groups) with headache, fatigue and muscle pain most frequently reported, table 2a. Only two participants, both in the low dose group, reported a fever post-vaccination. Three reactions reported by 2 participants were classified as severe, one participant with headache and fatigue and another reported fatigue, both participants recovered within 24 hours. All other local or systemic reactions were reported as mild or moderate (table 2b) and the majority resolved within 7 days of vaccination. Unsolicited adverse events, including laboratory abnormalities thought to be related to vaccination were reported in 27 (17·6%) participants, with 23·5%, 13·7% and 15·7% reported in the low, medium and high dose groups respectively. The most common laboratory abnormality was a rise in the red blood cell/erythrocyte sedimentation rate (ESR), which was reported in 11 (7·2%) of participants at a single centre. In addition, neutropenia was observed in 2 participants (1·3%) and eosinophilia (0·7%) and thrombocytopenia (0·7%) in one participant each up to Day 36. There were two cases of COVID-19, confirmed by nucleic acid amplification testing. One mild COVID-19 case occurred in a participant in the medium dose group, 16 days after the first vaccination. The other COVID-19 case was moderate in severity in a participant in the low dose group 4 days after the second vaccination. One AESI was reported which was a mild case of chilblains 4 days after first vaccination. The participant tested negative for COVID-19, had normal platelets and the event was considered unrelated to vaccination and a second vaccine dose administered without further event. As per protocol definition, any AESI was treated as serious adverse event in this study. However, no other AESI nor SAE was reported up to day 36.

**Table 2a.** Injection site reactions 7 days after any vaccine by frequency and severity.

| Adverse Event/Severity | Low Dose<br>(N=51)<br>n (%) | Medium<br>Dose<br>(N=51)<br>n (%) | High Dose<br>(N=51)<br>n (%) | Overall<br>(N=153)<br>n (%) |
| --- | --- | --- | --- | --- |
| Participants with at least one Solicited Injection Site Reaction | 35 (68.6) | 31 (60.8) | 36 (70.6) | 102 (66.7) |
| Mild (Grade 1) | 32 (62.7) | 27 (52.9) | 32 (62.7) | 91 (59.5) |
| Moderate (Grade 2) | 3 (5.9) | 4 (7.8) | 4 (7.8) | 11 (7.2) |
| Injection Site Tenderness | 32 (62.7) | 23 (45.1) | 34 (66.7) | 89 (58.2) |
| Mild (Grade 1) | 30 (58.8) | 20 (39.2) | 33 (64.7) | 83 (54.2) |
| Moderate (Grade 2) | 2 (3.9) | 3 (5.9) | 1 (2.0) | 6 (3.9) |
| Injection Site Pain | 19 (37.3) | 21 (41.2) | 24 (47.1) | 64 (41.8) |
| Mild (Grade 1) | 17 (33.3) | 20 (39.2) | 21 (41.2) | 58 (37.9) |
| Moderate (Grade 2) | 2 (3.9) | 1 (2.0) | 3 (5.9) | 6 (3.9) |
| Injection Site Itching | 3 (5.9) | 3 (5.9) | 2 (3.9) | 8 (5.2) |
| Mild (Grade 1) | 3 (5.9) | 3 (5.9) | 2 (3.9) | 8 (5.2) |
| Injection Site Swelling | 1 (2.0) | 0 | 1 (2.0) | 2 (1.3) |
| Mild (Grade 1) | 1 (2.0) | 0 | 0 | 1 (0.7) |
| Moderate (Grade 2) | 0 | 0 | 1 (2.0) | 1 (0.7) |

**Table 2b.** Solicited adverse reactions 7 days after any vaccine by frequency and severity.

| Adverse Event/Severity | Low Dose<br>(N=51)<br>n (%) | Medium<br>Dose<br>(N=51)<br>n (%) | High Dose<br>(N=51)<br>n (%) | Overall<br>(N=153)<br>n (%) |
| --- | --- | --- | --- | --- |
| Participants with at least one Solicited Systemic Reaction | 37 (72.5) | 32 (62.7) | 37 (72.5) | 106 (69.3) |
| Mild (Grade 1) | 26 (51.0) | 23 (45.1) | 26 (51.0) | 75 (49.0) |
| Moderate (Grade 2) | 11 (21.6) | 9 (17.6) | 9 (17.6) | 29 (19.0) |
| Severe (Grade 3) | 0 | 0 | 2 (3.9) | 2 (1.3) |
| Headache | 28 (54.9) | 17 (33.3) | 26 (51.0) | 71 (46.4) |
| Mild (Grade 1) | 22 (43.1) | 14 (27.5) | 20 (39.2) | 56 (36.6) |
| Moderate (Grade 2) | 6 (11.8) | 3 (5.9) | 5 (9.8) | 14 (9.2) |
| Severe (Grade 3) | 0 | 0 | 1 (2.0) | 1 (0.7) |
| Fatigue | 23 (45.1) | 15 (29.4) | 22 (43.1) | 60 (39.2) |
| Mild (Grade 1) | 16 (31.4) | 10 (19.6) | 15 (29.4) | 41 (26.8) |
| Moderate (Grade 2) | 7 (13.7) | 5 (9.8) | 5 (9.8) | 17 (11.1) |
| Severe (Grade 3) | 0 | 0 | 2 (3.9) | 2 (1.3) |
| Muscle Pain | 18 (35.3) | 15 (29.4) | 17 (33.3) | 50 (32.7) |
| Mild (Grade 1) | 15 (29.4) | 13 (25.5) | 16 (31.4) | 44 (28.8) |
| Moderate (Grade 2) | 3 (5.9) | 2 (3.9) | 1 (2.0) | 6 (3.9) |
| Nausea/Vomiting | 5 (9.8) | 7 (13.7) | 6 (11.8) | 18 (11.8) |
| Mild (Grade 1) | 5 (9.8) | 6 (11.8) | 5 (9.8) | 16 (10.5) |
| Moderate (Grade 2) | 0 | 1 (2.0) | 1 (2.0) | 2 (1.3) |
| Fever/Body Temperature (T > 38°C) | 2 (3.9) | 0 | 0 | 2 (1.3) |
| Mild (Grade 1) | 0 | 0 | 0 | 0 |
| Moderate (Grade 2) | 2 (3.9) | 0 | 0 | 2 (1.3) |

In the SARS-CoV-2 neutralising MNA_50_ assay, the highest median GMT recorded to date were reached on day 36 in all dose groups, in a dose dependent manner (high dose group, 520 [95% CI 421·49, 667·52]; medium dose, 222·3 [95% CI 171·84, 287·67]; low dose, 161·1 [CI 121·35, 213·82]) shown in table 3. The MNA_50_ GMT were significantly higher in the high dose group compared to both the low and medium dose groups (p < 0·001). The seroconversion rates of neutralising antibodies by day 36 were 90·0% (95% CI:78·0%·,97·0%) in the high dose group, 73·5% (95% CI: 59%,85%), in the medium dose group and 51% (95%CI: 37%,65%) in the low dose group.

**Table 3.** SARS-CoV-2 Neutralising Antibody Titres (ND40) at baseline prior to vaccination, 22 days after 1^st^ vaccination and 14 days after second vaccination;

| Visit |  | Low Dose<br>(N=51) | Medium Dose<br>(N=49) | High Dose<br>(N=50) | Overall<br>(N=150) |
| --- | --- | --- | --- | --- | --- |

|  |  | n (%) | n (%) | n (%) | n (%) |
| --- | --- | --- | --- | --- | --- |
| Day 1 |  | 51 | 49 | 50 | 150 |
|  | GMT (95% CI) | 29.0 (29.00, 29.00) | 30.7 (28.51, 33.15) | 30.0 (28.83, 31.23) | 29.9 (29.08, 30.73) |
|  | Min, Max | 29.0, 29.0 | 29.0, 232.0 | 29.0, 72.0 | 29.0, 232.0 |
|  | p-value (Overall Dose Groups Comparison) |  |  |  | 0.349 |
| Day 22 |  | 50 | 48 | 48 | 146 |
|  | GMT (95% CI) | 35.9 (32.31, 39.93) | 38.3 (31.92, 46.06) | 46.5 (38.79, 55.66) | 39.9 (36.45, 43.76) |
|  | Min, Max | 29.0, 115.0 | 29.0, 640.0 | 29.0, 359.0 | 29.0, 640.0 |
|  | p-value (Overall Dose Groups Comparison) |  |  |  | 0.089 |
| Day 36 |  | 51 | 48 | 50 | 149 |
|  | GMT (95% CI) | 161.1 (121.35, 213.82) | 222.3 (171.84, 287.67) | 530.4 (421.49, 667.52) | 266.6 (226.79, 313.34) |
|  | Min, Max | 29.0, 2171.0 | 29.0, 1307.0 | 29.0, 2033.0 | 29.0, 2171.0 |
|  | p-value (Overall Dose Groups Comparison) |  |  |  | <0.001 |
|  | p-value (Low Dose vs Medium dose) [2] |  |  |  | 0.204 |
|  | p-value (Medium Dose vs High Dose) [2] |  |  |  | <0.001 |
|  | p-value (Low Dose vs High Dose) |  |  |  | <0.001 |

GMT: Geometric Mean Titre, CI: Confidence Interval; Values below Limit of Quantification (58) are set to 29.0

A similar dose dependent response, with a peak response at day 36, was observed in anti-spike-IgG GMT measured by ELISA (high dose, 2147·9 [95% CI 1705·98, 2704·22]; medium dose, 691·6; [95% CI 494·.91, 966·.52]; low dose, 325; [95% CI 245·45, 430·36]), table 4. The anti-spike IgG GMT were significantly higher in the high dose groups in comparison to the other dose groups (p < 0.001). The neutralisation titres at day 36 correlated with anti-S-protein specific binding IgG (r=0·79 and p < 0·001), shown in figure 2. Median IFN-γ secreting ELISpot responses to Spike (S), Nucleocapsid (N) and Membrane (M) protein increased from baseline in all dose groups by day 36. In the high dose group, a reactive response was observed in 75% (34/45 participants) against S-protein,(figure 3), 36% (16/45)against M-protein and 49% (22/45) against N-protein,(supplementary material).

**Table 4.** IgG antibody Titres Against SARS-COV-2 S-protein at baseline prior to vaccination, 22 days after 1^st^ vaccination and 14 days after second vaccination (day 36)

| Visit |  | Low Dose<br>(N=51)<br>n (%) | Medium Dose<br>(N=49)<br>n (%) | High Dose<br>(N=50)<br>n (%) | Overall<br>(N=150)<br>n (%) |
| --- | --- | --- | --- | --- | --- |
| Day 1 |  | 51 | 49 | 50 | 150 |
|  | GMT (95% CI) | 25.6 (24.59,<br>26.69) | 26.4 (24.07,<br>29.05) | 25.0 (25.00,<br>25.00) | 25.7 (24.84,<br>26.54) |
|  | Min, Max | 25.0, 87.2 | 25.0, 390.8 | 25.0, 25.0 | 25.0, 390.8 |
|  | p-value (Overall Dose<br>Groups Comparison) |  |  |  | 0.604 |
| Day 22 |  | 51 | 49 | 50 | 150 |
|  | GMT (95% CI) | 27.1 (25.10,<br>29.16) | 31.9 (26.15,<br>38.86) | 30.0 (26.92,<br>33.48) | 29.6 (27.35,<br>31.93) |
|  | Min, Max | 25.0, 99.7 | 25.0, 1348.6 | 25.0, 165.0 | 25.0, 1348.6 |
|  | p-value (Overall Dose<br>Groups Comparison) |  |  |  | 0.293 |
| Day 36 |  | 51 | 49 | 50 | 150 |
|  | GMT (95% CI) | 325.1 (245.45,<br>430.46) | 691.6 (494.91,<br>966.52) | 2147.9<br>(1705.98,<br>2704.22) | 780.6 (643.85,<br>946.37) |
|  | Min, Max | 25.0, 5566.1 | 25.0, 8637.6 | 116.8, 15419.5 | 25.0, 15419.5 |
|  | p-value (Overall Dose<br>Groups Comparison) |  |  |  | <0.001 |
|  | p-value (Low Dose vs<br>Medium dose) |  |  |  | 0.004 |
|  | p-value (Medium Dose vs<br>High Dose) |  |  |  | <0.001 |
|  | p-value (Low Dose vs<br>High Dose) |  |  |  | <0.001 |

GMT: Geometric Mean Titre, CI: Confidence Interval, Values below Limit of Quantification (50.3) are set to 25.0

**Figure 2.**
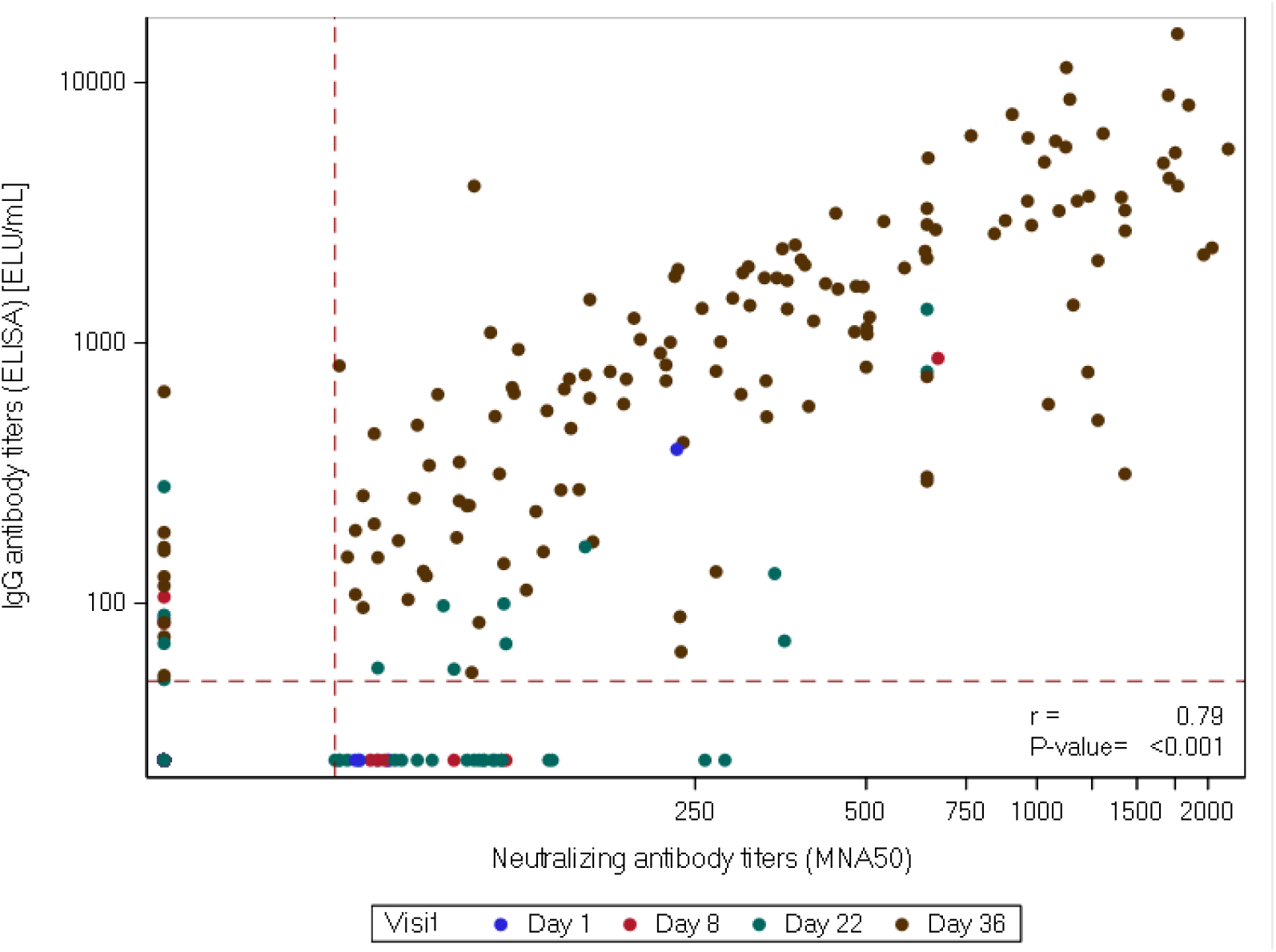
Correlation between neutralizing antibody titres ND50 and IgG antibody titres. Red dotted lines present the limit of detection for ELISA (50.3 ELU/mL) and MNA (ND50=58). (r) represents Pearson correlation coefficient

**Figure 3.**
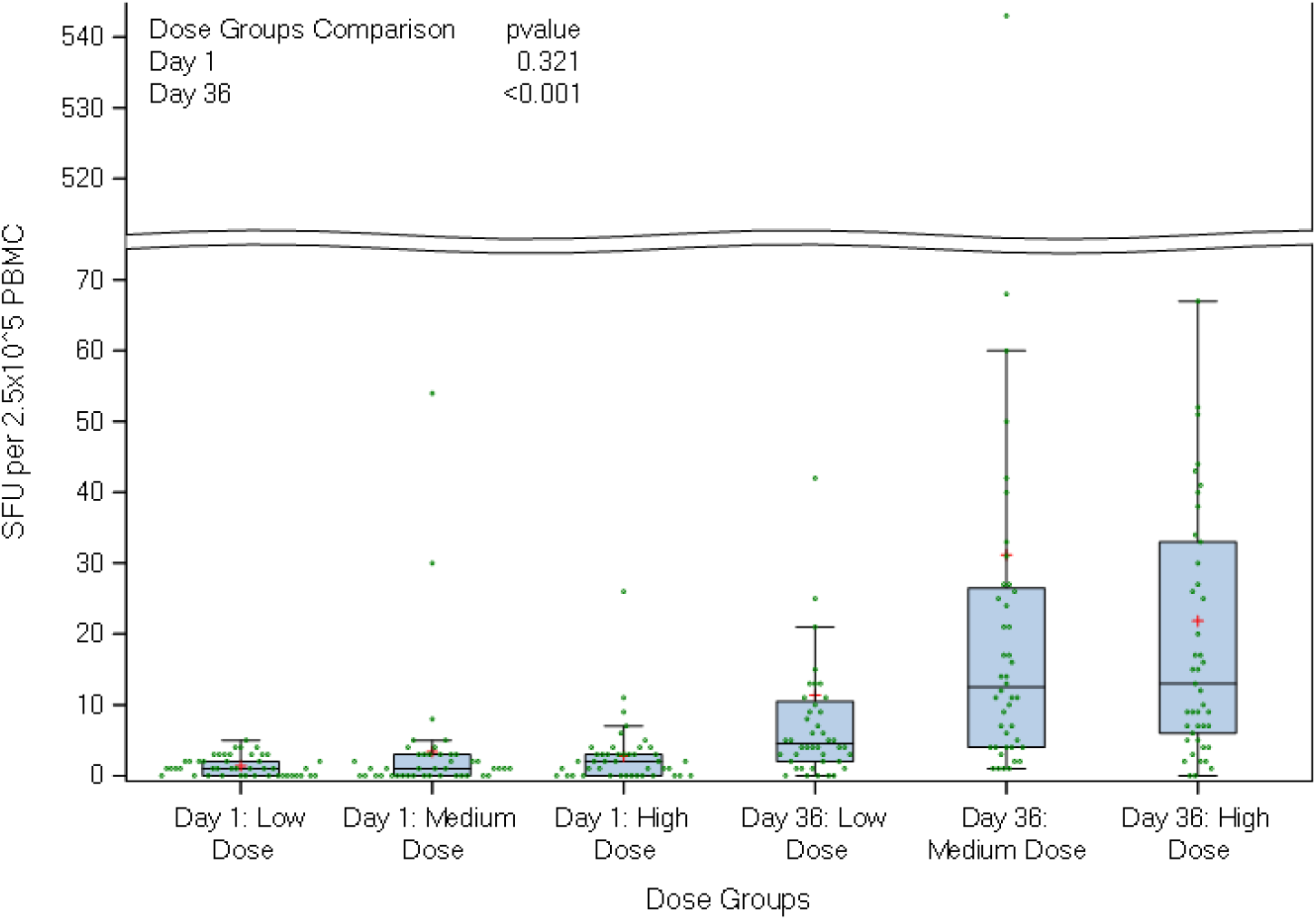
T-cell ELISpot responses to full sequence SARS-CoV-2 Spike protein. Boxplots show median, lower quartile and upper quartile; the horizontal line within each bar is the median and red + sign represents the mean value for each group. Green dots represent the measured values of SFU per 2.5×10^5 PBMC within each group

## Discussion

The vaccine candidate VLA2001 evaluated in this phase 1/2 study is a second wave vaccine candidate which includes the additional immune modulator CpG1018 to further broaden immune stimulation. The results from this study indicate that this vaccine candidate was able to induce both robust humoral and cellular immune responses. In general, inactivated vaccines adjuvanted with aluminium salts only induce minimal cellular immune responses and available data on SARS-CoV-2 vaccines of this kind demonstrate only a limited T-cell response.^4-7^ Zhang et al, reported poor T-cell responses to viral S-protein.^15^ In contrast, in a phase 1 study of BB152, a whole virion, inactivated vaccine formulated with a toll like receptor 7/8 agonist, CD4+ and CD8+ T-cell responses were detected in a subset of 16 participants.^7^

CpG 1018 is licensed as a vaccine adjuvant for a hepatitis B vaccine. The pre-clinical and clinical studies for that vaccine demonstrated that the addition of CpG 1018 increased antibody concentrations and resulted in robust T cell responses.^8^ Similarly, the results presented in this study show that the vaccine candidate VLA2001 was able to induce antigen specific T-cell responses against the S-protein, M and N protein, in many of the participants who received the high dose vaccine. Induction of both humoral and cellular immune responses against viral antigens beyond the S-protein may be beneficial for cross-protection against variants by broadening epitope coverage and immune effector mechanisms. ^12,13^

There was a clear dose dependent increase in both neutralising and spike protein binding antibodies with a high correlation between them. Ascending concentrations were recorded 2 weeks after the second dose of vaccine, with seroconversion for S-protein binding antibodies of up to 100%. Three weeks after a single dose only 6 (9%) of all participants in all dose groups showed seroconversion. Antibody concentrations after a single dose may have reached higher concentrations if measured at a longer interval after vaccination as vaccine-induced antibody concentrations have been reported to peak about 4 weeks after vaccination for other COVID-19 vaccines after each dose. ^4, 14^

After the initial pandemic response, reactogenicity and tolerability may become a more important feature of booster vaccines, particularly if they are to be used annually and a choice of vaccines is available, to help maintain uptake and compliance. VLA2001, given as a two-dose schedule with a 3-week interval was safe and well tolerated at all dose levels and no safety concerns were identified by an independent Data Safety Monitoring Board. Importantly, only two participants reported severe solicited AEs and there were no severe vaccine-related unsolicited events as well as no serious treatment-related adverse events. Of note, no cases of fever were reported in the two highest dose groups. Currently approved viral vector and mRNA vaccines have been reported to cause fever and chills in between 16% and 24% of individuals depending on the age and dose (first or second). ^15,16^ Furthermore, using the same approach as used for the proven and well-established annual modification of inactivated influenza vaccines, an inactivated vaccine could be particularly well suited for use as an annual booster vaccine.

Our study has several limitations. Firstly, the population studied was young, healthy adults, who typically have higher immune responses post vaccination than older adults. While this is a classical approach in Phase 1/2, it will be necessary and important to include an older population in future trials. Secondly, we did not study the response to immunization in subjects seropositive for SARS-CoV-2, so we cannot yet quantify any safety issues or a potential booster effect of the first dose that might be present in such individuals, as has been seen for other vaccines Thirdly, we have only assessed antibody titres at 2 weeks after the second dose at the moment. Further follow-up of participants is ongoing to provide immunogenicity data from time points later after the primary series.

The highest dose group has been selected for further clinical development based on comparable safety and superior immunogenicity results. At the time of this publication, the sponsor has initiated a comparative, immunogenicity Phase 3 trial that will aim to predict vaccine efficacy by employing a primary endpoint of superiority of the GMT ratio of neutralizing antibody titres following 2 doses of VLA2001 compared to 2 doses of AZD1222.^17^

## Data Availability

Data available on request

## Contributors

AF is the chief investigator. RL, CD, SF and CG were the study site principal investigators. RL and CT prepared this report. All authors critically reviewed and approved the final version.

## Data sharing

The study protocol is provided in the appendix.

## Acknowledgements

This study was funded by the UK Government and the Vaccine Task Force. This research was supported by the National Institute for Health Research. The views expressed are those of the author(s) and not necessarily those of the NIHR or the Department of Health and Social Care. The investigators express their gratitude for the contribution of all the trial participants.

## Research in context

### Evidence before this study

We searched Pubmed for research articles published from database inception until April 13^th^ 2021, with no language restrictions using the terms “inactivated”, “SARS-CoV-2”, “vaccine”, AND “clinical trial”. There are studies in preprint which have not been included in this evidence review. We identified clinical trial data on six other inactivated SARS-CoV-2 vaccines. A dose-escalation study of a whole virion inactivated vaccine made using a SARS-CoV-” (CN02) strain vaccine, adsorbed onto aluminium hydroxide (Coronavac, Sinovac Life Sciences, Beijing), was tested in health adults aged 18 to 59 in a two-dose regimen. seroconversion rates of neutralising antibody were reported as 92% and 98%, the 3µg and 6µg groups, respectively. T-cell responses, as measured by T-cell ELISpot were low. The overall incidence of adverse reactions was 29%. BBIBP-CorV (Sinopharm, Beijing), a whole virion inactivated vaccine with aluminium hydroxide adjuvant was tested with 3 different concentrations and as a single or two dose regimen with varying dose intervals. The intermediate dose (4 µg) given on days 0 and 21 or 0 and 28 was found to produce higher neutralisation titres than a single 8 µg dose or a dose administered on days 0 and 14. Up to 46% of participants experienced an adverse reaction in the first 7 days after vaccination. No T-cell responses formally reported but authors report that preliminary results suggest minimal evidence of T-cell activation. An interim analysis of an inactivated vaccine with aluminium of 2 randomised control trials that included 320 participants who received one of the three doses as either a 3-dose schedule (days 1, 28 and 56) or a two-dose schedule (days 0 and 14 or days 0 and 21). The vaccine was shown to produce higher neutralising antibodies when given at 21 or 28 days compared to 14 days and a third dose resulted in further increase in neutralising antibody titres. The incidence of adverse reactions was reported as 15%. No discernible T-cell response was noted through the measurement of T-cell subsets or the evaluation of cytokine profiles. A phase 1, investigating a vaccine alum adsorbed, inactivated using a patented two step deactivation process was evaluated as a 2-dose regimen given at an interval of either 14 or 28 days at three different dose ranges (Institute of Medical Biology and Chinese Academy of Medical Sciences). Adverse reactions were reported in 4.2% to 33.3% of participants depending on dose group and interval. Seroconversion defined as a fourfold increase in neutralising antibody concentration after vaccination was between 54.2% to 100%. A phase 1 / 2 trial of an inactivated, alum adsorbed KCONVAC vaccine was initiated with dose escalation from 5 to 10ug in 60 healthy participants with both the 5µg and 10µg doses used in the phase 2. In both phases the vaccines were administered as a two-dose regimen administered either 14 days or 28 days apart. Seroconversion was reported in 83% to 100% in the 5µg and 10µg dose group respectively, across both phases of the study. Reactive T-cell IFN-γ-ELISpot were reported in 57% to 63% in the 5µg and 10µg dose group, respectively, 14 days after the first dose of vaccine. Finally, BBV152, an alum-adjuvanted inactivated vaccine combined with a TL7/8 agonist, given as a 2-dose regiment, 2 weeks apart and two different doses with and without the TL7/8 agonist and an aluminium alone group. The rates of seroconversion ranged between 82.8% and 87.9% with no significant difference in the groups who received the TL7/8 agonist containing vaccine. In contrast, in small subsets (13 participants) in each vaccine group cellular responses were observed in the group receiving the TLR 7/8 containing vaccine but not in the vaccines without this. There were no safety concerns reported in any of these trials. Peer reviewed data on efficacy is limited however a recent manuscript of a randomised clinical trial including two alum-adjuvanted, inactivated vaccines, which demonstrated an efficacy between 72.8% and 78.1% against laboratory-confirmed symptomatic COVID-19 disease. A prospective, observation study has also recently demonstrated the effectiveness of CoronaVAC as 65.9% against laboratory confirmed disease and 87.5% against hospitalisation.

### Added value of this study

This will be the seventh published study of a vaccine against SARS-CoV-2 which has been tested in healthy adults aged 18 to 55 and the first using the TLR9 agonist, CpG. The vaccine was shown to be safe and well tolerated. Neutralising and binding antibodies against SARS-CoV-2 were induced at all doses evaluated after 2 doses of vaccine and increased in a dose dependent manner. Cellular immune responses were also demonstrated by 14 days after the second dose of vaccine. The cellular response is believed to be stimulated by the addition of the CpG adjuvant, a Toll like receptor 9 agonist.

### Implications of all the available evidence

Inactivated vaccines against SARS-CoV-2 contribute to the portfolio of COVID-19 vaccines that are required to ensure a resilient global vaccine supply. An important limitation of inactivated vaccines is can be weak cellular immunity however the addition of suitable adjuvants such as CpG may overcome this limitation. Further assessment of the efficacy and immunogenicity of this vaccine in relation to an approved comparator vaccine is under way.

|  |  |
| --- | --- |
| Sarah E. Kidd | University Hospitals Bristol and Weston NHS Foundation Trust, Bristol, UK |
| Kelly L. Turner | University Hospitals Bristol and Weston NHS Foundation Trust, Bristol, UK |
| Kirsty M. Phillips | University Hospitals Bristol and Weston NHS Foundation Trust, Bristol, UK |
| Oliver J Griffiths | University Hospitals Bristol and Weston NHS Foundation Trust, Bristol, UK |
| Julie Cloake | University Hospitals Bristol and Weston NHS Foundation Trust, Bristol, UK |
| Agnieszka Skorko | University Hospitals Bristol and Weston NHS Foundation Trust, Bristol, UK |
| Amisha Desai | NIHR/Wellcome Trust Clinical Research Facility, University Hospitals Birmingham NHS Foundation Trust |
| Simon D. Horley | NIHR/Wellcome Trust Clinical Research Facility, University Hospitals Birmingham NHS Foundation Trust |
| Ehsaan EQ Qureshi | NIHR/Wellcome Trust Clinical Research Facility, University Hospitals Birmingham NHS Foundation Trust |
| Tom T Parker | NIHR/Wellcome Trust Clinical Research Facility, University Hospitals Birmingham NHS Foundation Trust |
| Helen J Smith | NIHR/Wellcome Trust Clinical Research Facility, University Hospitals Birmingham NHS Foundation Trust |
| Daniel R. Owens | NIHR Southampton Clinical Research Facility and Biomedical Research Centre, University Hospital Southampton NHS Foundation Trust, Southampton, UK; Faculty of Medicine and Institute for Life Sciences, University of Southampton, Southampton, UK |
| Christopher J. Edwards | NIHR Southampton Clinical Research Facility and Biomedical Research Centre, University Hospital Southampton NHS Foundation Trust, Southampton, UK; Faculty of Medicine and Institute for Life Sciences, University of Southampton, Southampton, UK |
| Katrina Cathie | NIHR Southampton Clinical Research Facility and Biomedical Research Centre, University Hospital Southampton NHS Foundation Trust, Southampton, UK; Faculty of Medicine and Institute for Life Sciences, University of Southampton, Southampton, UK |
| Matthew Stokes | NIHR Southampton Clinical Research Facility and Biomedical Research Centre, University Hospital Southampton NHS Foundation Trust, Southampton, UK; Faculty of Medicine and Institute for Life Sciences, University of Southampton, Southampton, UK |
| Stephen Saich | NIHR Southampton Clinical Research Facility and Biomedical Research Centre, University Hospital Southampton NHS Foundation Trust, Southampton, UK; Faculty of Medicine and Institute for Life Sciences, University of Southampton, Southampton, UK |
| Lisa Berry | NIHR Southampton Clinical Research Facility and Biomedical Research Centre, University Hospital Southampton NHS Foundation Trust, Southampton, UK; Faculty of Medicine and Institute for Life Sciences, University of Southampton, Southampton, UK |
| Laura Presland | NIHR Southampton Clinical Research Facility and Biomedical Research Centre, University Hospital Southampton NHS Foundation Trust, Southampton, UK; Faculty of Medicine and Institute for Life Sciences, University of Southampton, Southampton, UK |
| Louise Haskell | NIHR Southampton Clinical Research Facility and Biomedical Research Centre, University Hospital Southampton NHS Foundation Trust, Southampton, UK; Faculty of Medicine and Institute for Life Sciences, University of Southampton, Southampton, UK |
| Christopher J Edwards | NIHR Southampton Clinical Research Facility and Biomedical Research Centre, University Hospital Southampton NHS Foundation Trust, Southampton, UK; Faculty of Medicine and Institute for Life Sciences, University of Southampton, Southampton, UK |
| Andrew Barr | The Newcastle upon Tyne Hospitals NHS Foundation Trust, Newcastle upon Tyne, NE1 4LP, UK |
| Zoe Kirkham | The Newcastle upon Tyne Hospitals NHS Foundation Trust, Newcastle upon Tyne, NE1 4LP, UK<br>Translational and Clinical Research Institute, Newcastle University, |
|  | Newcastle upon Tyne, NE2 4HH, UK |
| Jade Deighton | The Newcastle upon Tyne Hospitals NHS Foundation Trust,<br>Newcastle upon Tyne, NE1 4LP, UK |
| Adam Scadeng | Newcastle NIHR Clinical Research Facility, Newcastle upon Tyne,<br>NE1 4LP, UK |
| Jennifer Bainbridge | Newcastle NIHR Clinical Research Facility, Newcastle upon Tyne,<br>NE1 4LP, UK |
| Lucie Robinson | Newcastle NIHR Clinical Research Facility, Newcastle upon Tyne,<br>NE1 4LP, UK |
| Mark Walker | The Newcastle upon Tyne Hospitals NHS Foundation Trust,<br>Newcastle upon Tyne, NE1 4LP, UK; Translational and Clinical<br>Research Institute, Newcastle University, Newcastle upon Tyne,<br>NE2 4HH, UK; Newcastle NIHR Clinical Research Facility,<br>Newcastle upon Tyne, NE1 4LP, UK |

## Supplementary material

**Supplementary figure 1.**
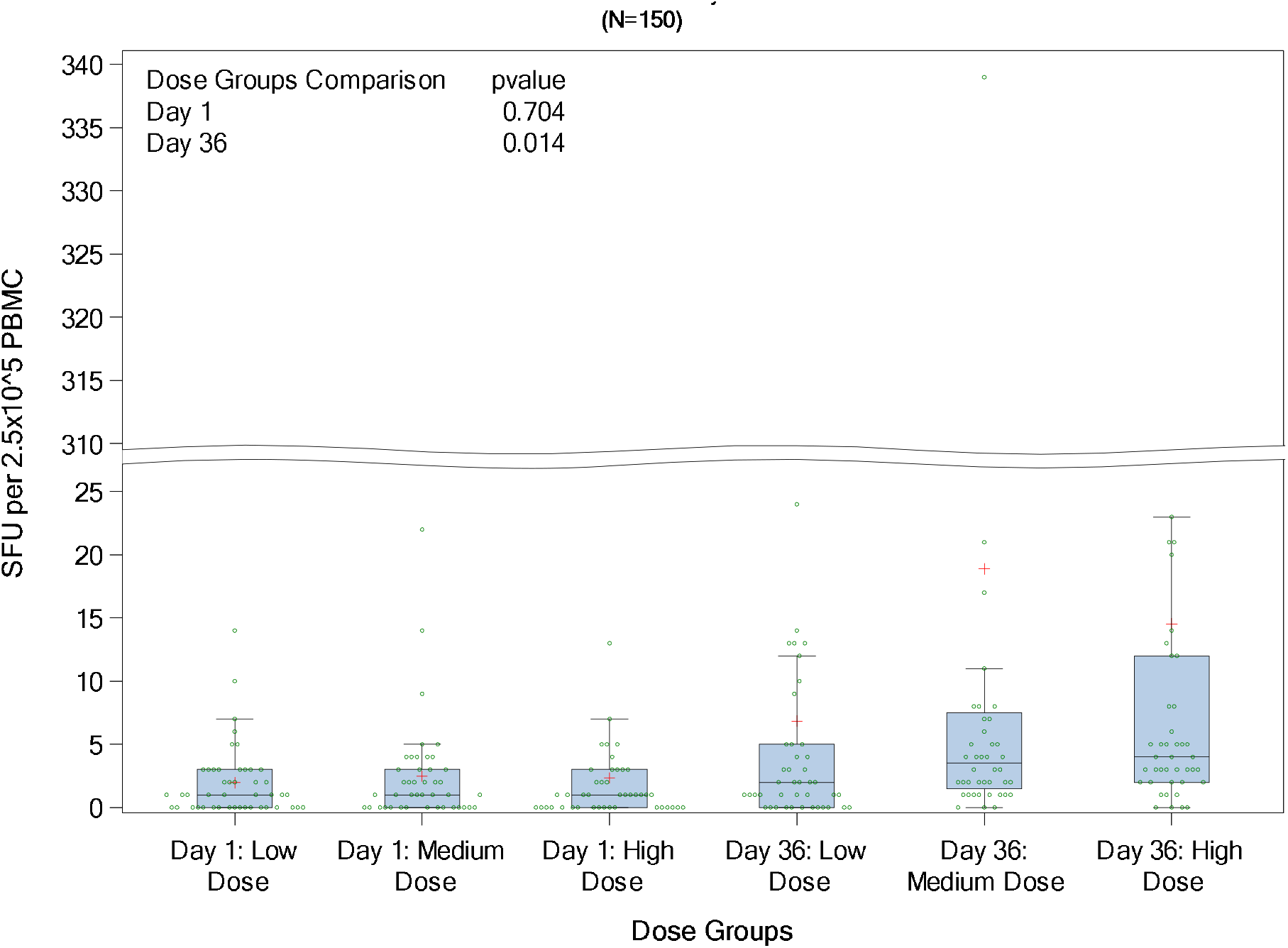
T-cell ELISpot responses to SARS-CoV-2 Membrane protein. Boxplots show median, lower quartile and upper quartile; the horizontal line within each bar is the median and red + sign represents the mean value for each group. Green dots represent the measured values of SFU per 2.5×10^5 PBMC within each group

**Supplementary figure 2.**
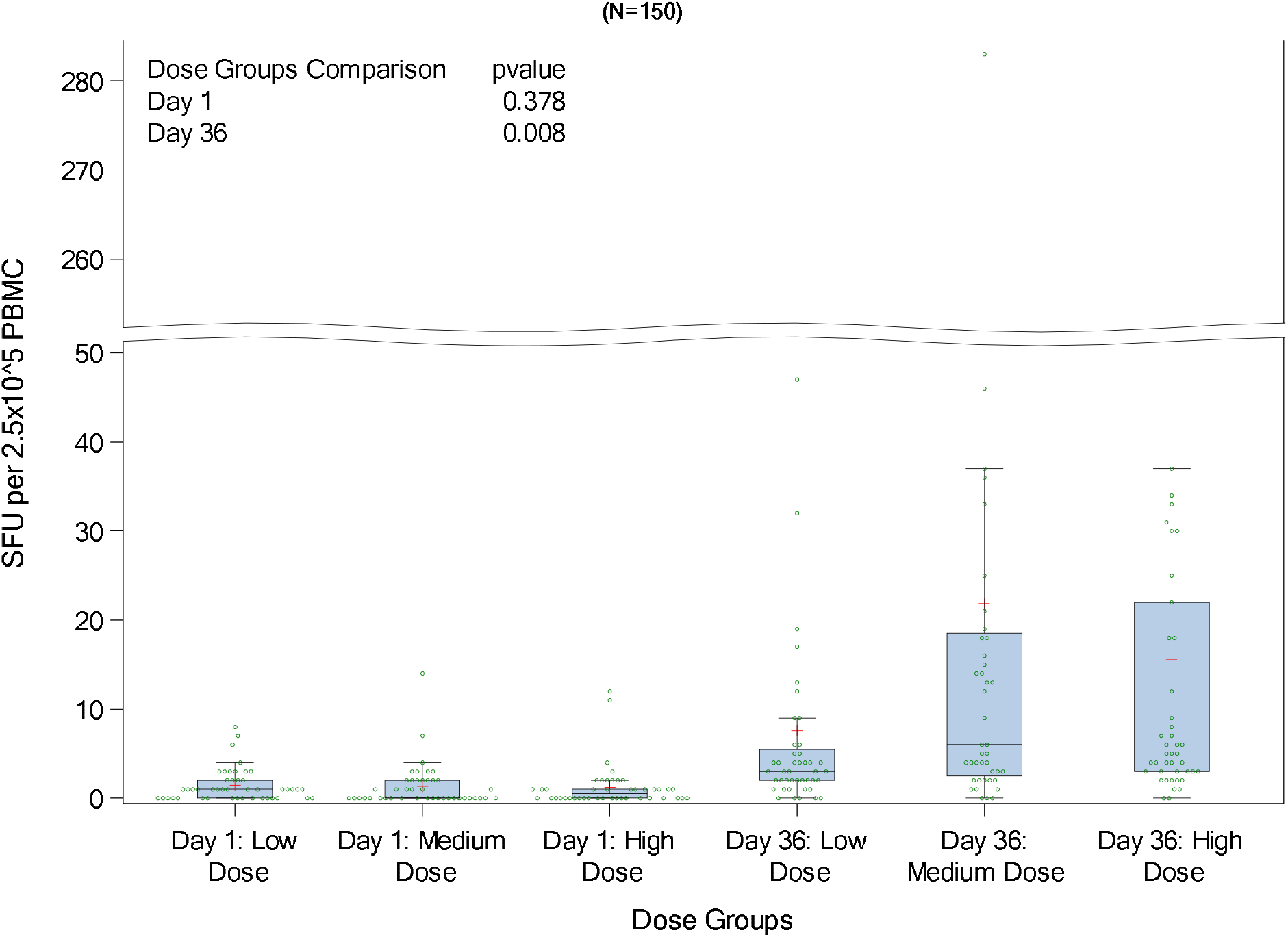
T-cell ELISpot responses to SARS-CoV-2 Nucleocapsid protein. Boxplots show median, lower quartile and upper quartile; the horizontal line within each bar is the median and red + sign represents the mean value for each group. Green dots represent the measured values of SFU per 2.5×10^5 PBMC within each group

## Appendix A Study Protocol

